# Prevalence, incidence and risk factors for myopia among urban and rural children in southern China: protocol for a school-based cohort study

**DOI:** 10.1101/2021.03.26.21254308

**Authors:** Xin Chen, Guofang Ye, Yuxin Zhong, Ling Jin, Xiaoling Liang, Yangfa Zeng, Yingfeng Zheng, Ian Morgan, Yizhi Liu

**Affiliations:** State Key Laboratory of Ophthalmology, Zhongshan Ophthalmic Center, Sun Yat-sen University, Guangzhou, China; Guangzhou Regenerative Medicine and Health Guangdong Laboratory, Guangzhou, China; Research Units of Ocular Development and Regeneration, Chinese Academy of Medical Sciences, China; Research School of Biology, Australian National University, Canberra, Australia

**Keywords:** Incidence, Prevalence, Myopia, Rural, Urban, China

## Abstract

**Introduction:** Myopia is the common cause of reduced uncorrected visual acuity among school-age children. It is more prevalent in urban than in rural areas. Although many myopia studies have focused on the effect of urbanization, it remains unclear how visual experience in urban regions could affect childhood myopia. This study aims to investigate the incidence and prevalence of myopia among school-age children in urban and rural settings, thereby identifying the environmental factors that affect the onset and progression of myopia.

**Methods and analysis:** A school-based cohort study will be conducted. We will enroll all first-grade students from an urban (10 primary schools) and a rural (10 primary schools) regions of Zhaoqing City, China. Over the 3-year follow-up period, students will receive detailed eye examinations annually and complete questionnaires about living habits and environment. In a 5% random subsample of the cohort, physical activity, light intensity, and eye-tracking data will be obtained using wearable devices. The primary outcome is incident myopia, defined as myopia detected during follow up among those without myopia at baseline.

**Ethics and Dissemination:** Ethics approval was obtained from the ethics committee of the Zhongshan Ophthalmic Center (number: 2019KYPJ171). Study findings will be published in a peer-reviewed journal.

**Trial registration number:** NCT04219228

**ARTICLE SUMMARY:** *Strengths and limitations of this study:*

- This is a longitudinal, school-based cohort study to assess the incidence and prevalence of myopia among school-age children in both urban and rural settings.
- Large sample size and the representative study population.
- Light exposure and daily activities will be measured using wearable devices in a random subsample of the cohort.
- A myopia prediction model will be established using a deep learning algorithm based on the collected data.

## INTRODUCTION

Myopia is the major cause of reduced uncorrected visual acuity among children and adolescents, particularly in the urban areas of East Asia.^1^ A higher prevalence rate of myopia has been consistently reported in urban than in rural regions.^2^ The effects of urbanization may be confounded by factors such as education, socioeconomic status, and outdoor exposure. It is still not entirely clear whether and how visual experience in an urban environment could have an impact on the development and progression of myopia.^3^

There is now considerable evidence that increased time outdoors reduces the onset of myopia, ranging from cross-sectional and longitudinal epidemiological studies,^4-5^ through to randomized clinical trials.^6-8^ In addition, a proposed mechanism based on exposures to brighter light outdoors has been demonstrated in animal models.^9-12^ However, not all studies have reported positive effects.^13^ Possible factors that could obscure a real association include lack of variation in time outdoors, or in prevalence and incidence of myopia, poor measurement of time outdoors^14-16^ and myopia, and small sample size.

We therefore intend to conduct a school-based cohort study in both urban and rural settings to identify the incidence, prevalence and environmental risk factors of myopia among school-aged children. All participants will be enrolled from primary schools in Zhaoqing city, China, and followed up for three years. Our study design overcomes the above-mentioned factors by using the gold standard of cycloplegic refraction and objective measures of time outdoors on a sub-sample, on a large sample size recruited from a location that covers urban and rural areas where preliminary data indicate that the prevalence and incidence of myopia varies significantly.

## METHODS

The trial was registered on Clinicaltrials.gov (Identifier: NCT04219228). It is estimated that the study dates would be from December 14, 2019 to February 2, 2023.

### Objective

The purpose of this trial is to identify the incidence, prevalence and risk factors of myopia among school-age children in urban and rural regions of southern China.

### Study design

We will conduct a 3-year, school-based, longitudinal cohort study. This study adheres to the Declaration of Helsinki and ethics approval will be obtained from the Zhongshan Ophthalmic Center Institutional Review Board.

### Eligibility criteria

Inclusion Criteria:

- All first-grade students in urban (10 primary schools) and rural regions (10 primary schools), Zhaoqing city.

Exclusion Criteria:

- No.

### Trial setting and participants

Zhaoqing is one of the major cities in Guangdong province, and has a population of 4,084,600 (National Census 2016). Zhaoqing is chosen because of its relatively stable population covering a broad socioeconomic spectrum. To determine how environment affects the onset of myopia, first-grade students aged 6 to 7 will be recruited from both urban (Duanzhou district) and rural regions (Huaiji county). This period precedes the development of myopia in most children. The results on this sample will be compared with those from previous studies (e.g., the Sydney Myopia Study)^15^. **Table 1** shows the socio-demographic variables in different counties of Zhaoqing city. Satellite images for the Huaiji county and Duanzhou district are shown in **Figure 1**.

**Table 1.** Socio-demographic data in Zhaoqing City#.

| County | Number of inhabitants (thousands) | Land area (sq.km) | Population density (inhab. Per sq.km) | GDP in 2019 (billion RMB) |
| --- | --- | --- | --- | --- |
| **Huaiji\*** | **860** | **3,573** | **241** | **23.0** |
| Gaoyao | 800 | 2,071 | 386 | 42.0 |
| **Duanzhou\*** | **510** | **152** | **3,355** | **42.0** |
| Sihui | 500 | 1,258 | 397 | 58.7 |
| Guangning | 450 | 2,459 | 183 | 15.9 |
| Fengkai | 420 | 2,723 | 154 | 15.0 |
| Deqing | 360 | 2,258 | 159 | 15.0 |
| Dinghu | 190 | 512 | 371 | 12.6 |

#Data are available from The Guangdong Statistical Yearbook 2020.
\*Huaiji and Duanzhou districts will be selected as the rural and urban study areas, respectively.

**Figure 1.**
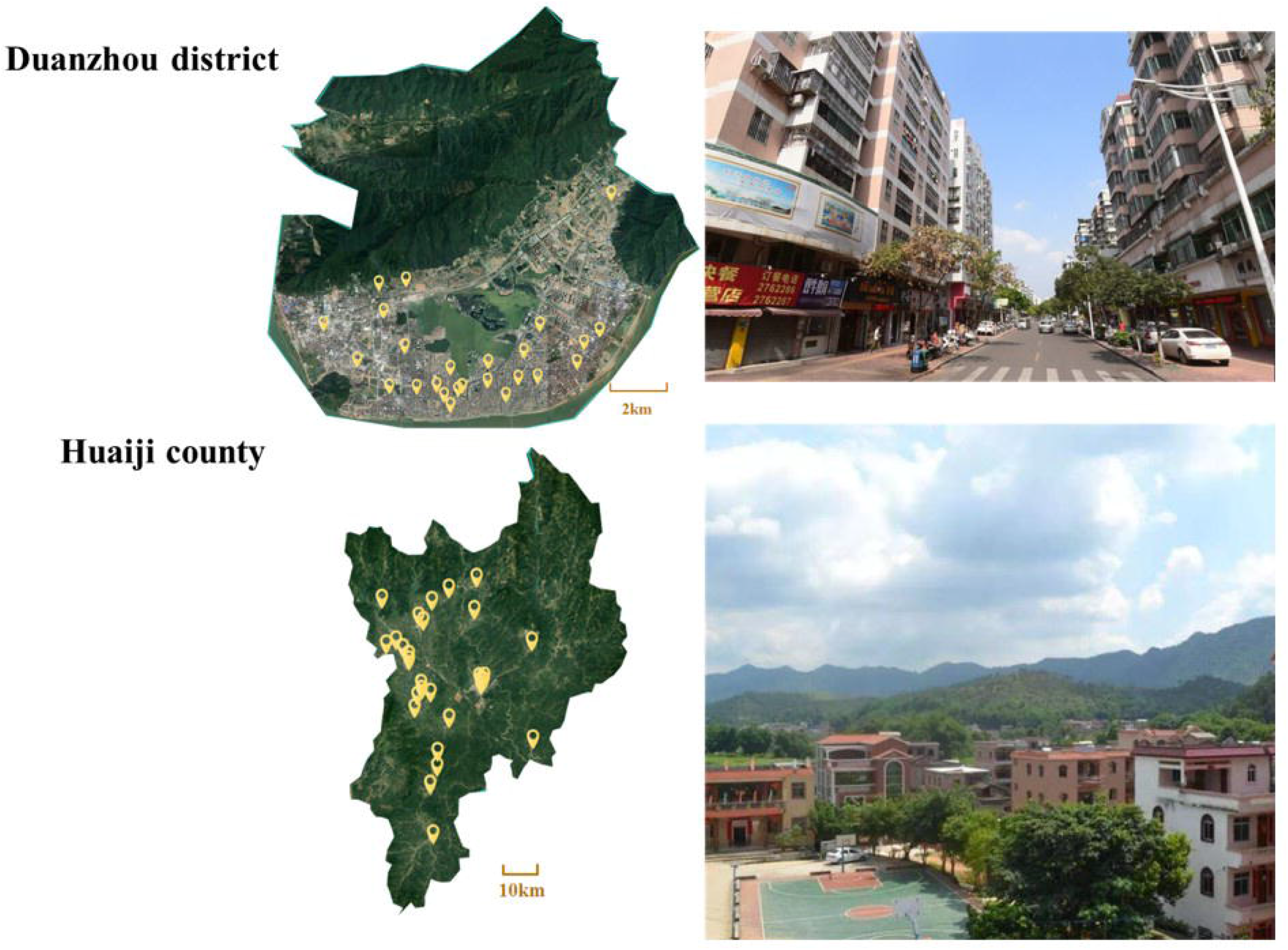
Satellite images for the Huaiji county and Duanzhou district. Satellite images for the Huaiji county and Duanzhou district. Most of the Duanzhou district (satellite image scale: 1:53858) are covered by residential buildings, whereas Huaiji county (satellite image scale: 1:433701) is mainly covered in greenery with mountains. Satellite images are available from: http://www.bigemap.com/source/tree/satel-244.html.

### Recruitment

A database containing annual examination result of uncorrected visual acuity (UCVA) of students was provided by Zhaoqing Education Bureau. There are 25 primary schools in Duanzhou district and 30 in Huaiji county. Given that most of the primary schools have more than 2 classes in each grade, we will exclude schools with less than 2 classes to keep an equal number of participants in each school. To ensure that all schools will be represented in the selected sample, all primary schools in Duanzhou District and Huaiji County will be stratified into 5 strata respectively based on previous results of visual acuity (uncorrected visual acuity ≥ 20/25). We will randomly select two schools from each stratum for our trial. In total, 10 schools from Duanzhou District and 10 from Huaiji County will be involved. Independent researchers will complete the randomization process and communicate the results to the research team. After that,research team will contact schools to recruit.

## SAMPLE SIZE

The size of each district group is about 200 persons, that is, about the number of students in the 1st grade of each school. Assuming the intraclass correlation coefficient (ICC) is 0.01, an estimate of the school’s accurate matching permissible expansion coefficient (IF) is 0.01 based on the visual acuity data. Based on an annual incidence of about 10 percent of myopia, an estimated 5% loss to follow-up rate due to transfer or drop-out is included, for a total of about 2,000 students, which will provide sufficient capacity to detect an approximately 50% difference in the incidence compared with the control group.

## OUTCOME MEASURES

### Primary outcome

The primary outcome is incident myopia, defined as myopia detected during follow up among those without myopia at baseline. Myopia in this study is defined as the right eye’s spherical equivalent refractive error (SER, sphere + 1/2 cylinder) of at least -0.5 diopters (D).

### Secondary outcomes

1. Prevalence of myopia [Time Frame: baseline] Myopia is defined as any eye’s SER (sphere + 1/2 cylinder) of at least -0.5 diopters (D).
2. Change in axial length [Time Frame: 1 year, 2 years, 3 years] Axial length will be measured with a non-contact optical device.
3. Prevalence of amblyopia, strabismus and other ocular abnormalities [Time Frame: baseline] Cover-uncover tests will be performed to detect strabismus. Any ocular abnormalities, including corneal opacities, lens opacities, and retinal diseases will be recorded based on slit lamp, direct ophthalmoscopic and/or mobile phone video examination. Participants with an uncorrected visual acuity 6/7.5 or worse with undergo subjective refraction to identify amblyopia.
4. Area under the receiver operating characteristic curve of the deep learning algorithm for the prediction of incident myopia [Time Frame: 1 year] The investigators will estimate the area under the receiver operating characteristic curve of the deep learning algorithm for the prediction of incident myopia.
5. Sensitivity and specificity of the deep learning algorithm for the prediction of incident myopia [Time Frame: 1 year] The investigators will estimate sensitivity and specificity of the deep learning algorithm for the prediction of incident myopia.
6. Area under the receiver operating characteristic curve of the deep learning algorithm for the prediction of fast progressing myope [Time Frame: 1 year] The investigators will estimate the area under the receiver operating characteristic curve of the deep learning algorithm for the prediction of fast progressing myope (a change in SER of 0.75D or more per year).
7. Sensitivity and specificity of the deep learning algorithm for the prediction of fast progressing myope [Time Frame: 1 year] The investigators will estimate the sensitivity and specificity of the deep learning algorithm for the prediction of fast progressing myope. Cycloplegic spherical refraction changes measured by an auto-refractometer will be used as the indicator of myopia progression.
8. Area under the receiver operating characteristic curve of the diagnostic algorithm in identifying abnormal vision screening result [Time Frame: baseline] The investigators will estimate the area under the receiver operating characteristic curve of the diagnostic algorithm in identifying abnormal vision screening result (e.g., abnormal eye lid, abnormal cornea, and strabismus detected with mobile devices).
9. Sensitivity and specificity of the diagnostic algorithm in identifying abnormal vision screening result [Time Frame: baseline] The investigators will estimate the sensitivity and specificity of the diagnostic algorithm in identifying abnormal vision screening result (e.g., abnormal eye lid, abnormal cornea, and strabismus detected with mobile devices).
10. Post-vision screening referral uptake [Time Frame: 3 months] Any referral uptake will be confirmed by telephone follow-up.

## EXAMINATIONS

The children’s height will be measured using a wall-mounted scale reading device. Weight will be recorded in kg to one decimal place. Ophthalmic examinations will include visual acuity, cover test and ocular dominance, noncycloplegic autorefraction, cycloplegia, ocular biometric measurements, cycloplegic auto-refraction, subjective refraction, and anterior and posterior segment examination. Any ocular abnormalities, including corneal opacities, lens opacities, and retinal diseases will be recorded based on slit lamp and direct ophthalmoscopic examination.

## QUESTIONNAIRES

A family questionnaire^5^ regarding educational attainment, occupations, refractive status, and history of eye disease will be administered only at baseline. The grade leader teacher or headteacher of each class will be asked to complete a class curriculum questionnaire^5^ about the curriculum schedule (e.g., school days, school holidays). Sleep quality and quantity will be measured with the PROMIS-Sleep Disorder questionnaire.^17^ Time (hour) spent in different activities (separately for outdoor activities, near work activities, screen time (separately for smartphone, tablet, TV, computer, games console)) will be measured with a Self-reported Previous Day Activity Recall Questionnaire. An OCT Instrument (Moptim Mocean-3500) will be used to obtain high-resolution macular images only at baseline.

## SUBSAMPLE STUDY

1. Spectacle-frame mounted distance sensor and eye-tracking Time (hour) spent in different activities will be measured with an eye tracker [Time frame: four randomly selected occasions each year, during the 3-year follow up]. On each occasion, participants will be asked to meet the study investigators at school, and they will be provided with a spectacle-frame eye tracker to be worn for 2 hours during the daytime and 2 hours during the night-time on the next day.
2. Wearable activity tracker Physical activity and light intensity will be measured with a wearable activity tracker [Time frame: four randomly selected occasions each year, during the 3-year follow up]. On each occasion, participants will be asked to meet the study investigators at school, and they will be provided with a wearable activity tracker to be worn on the wrist for 24 hours on the next day.

## WORKFLOW

The workflow will follow the order as above mentioned, starting from body measurement and ending with direct ophthalmoscopic examination (**Figures 2 and 3**). Participants will be followed up to maximize participation in the eye examinations at baseline and annually at years 1, 2 and 3, with the same examiner using the same set of instruments with regular calibration.

**Figure 2.**
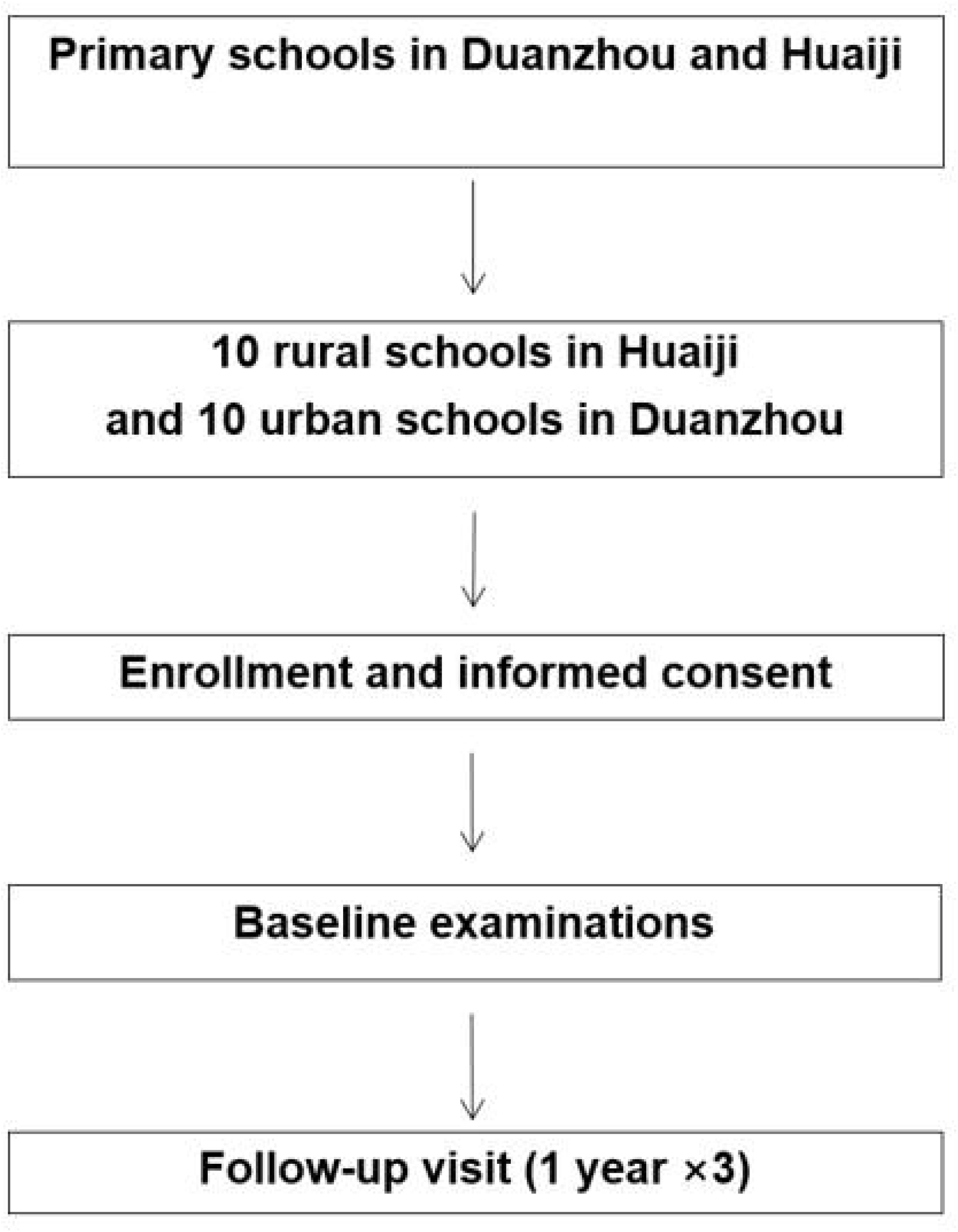
Flowchart of the trial.

**Figure 3.**
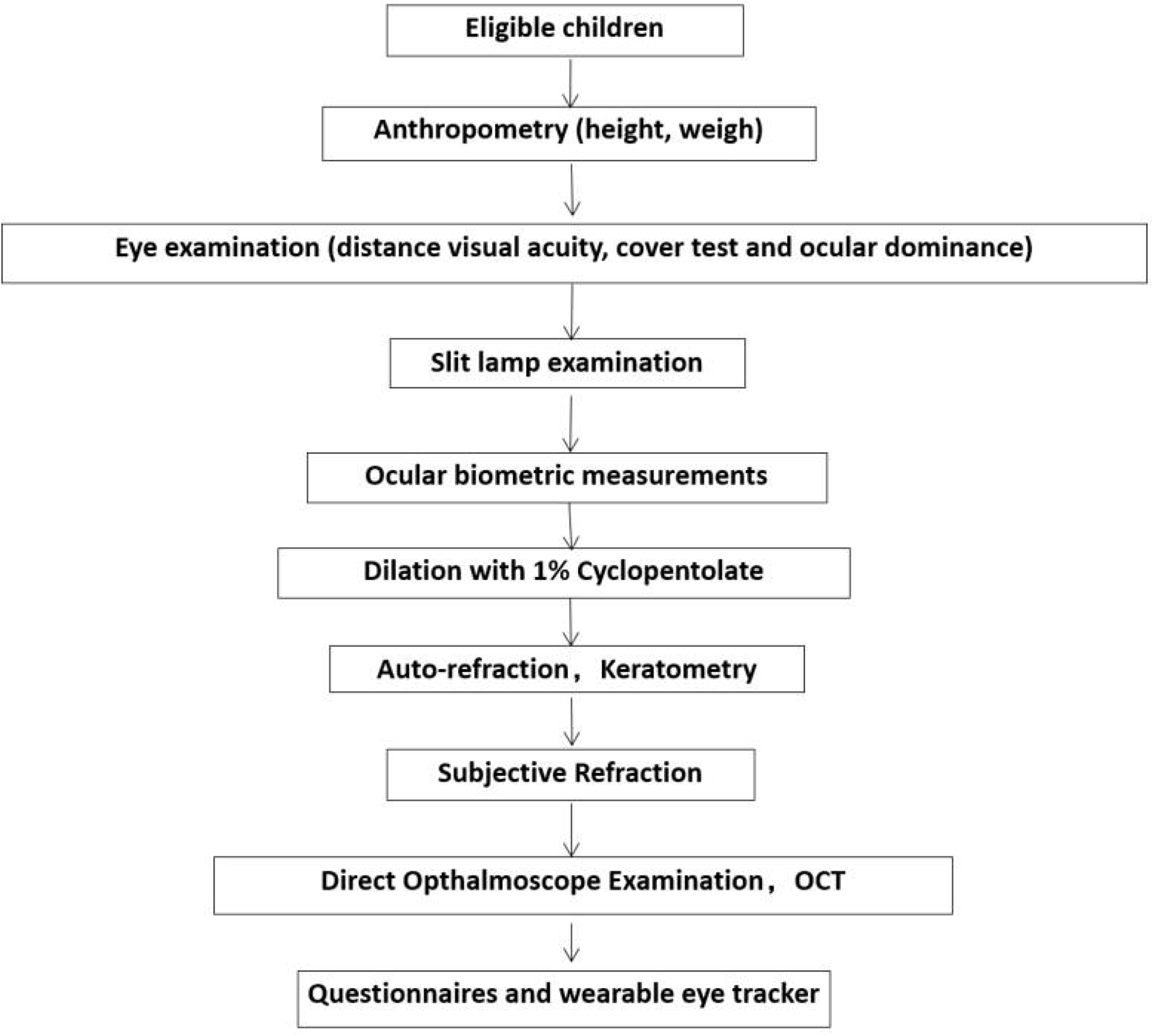
Flowchart of field examinations.

## DATA MANAGEMENT

The clinical examiner will regularly check the informed consent and eligibility for each participant to guarantee that all CRF forms are correct and in accordance with the original data. All errors and omissions will be noted, and where possible corrected. The examiner will also ensure every participant’s withdrawal and loss is recorded and explained in CRF, and all adverse events are recorded.

Results of eye examinations will be recorded on paper, except for those obtained from electronic questionnaires and ocular biometry measurements. To ensure data accuracy, data will be entered by two independent personnel for further data checking. After double data entry, we will select 5% of cases at random to identify any inconsistency between the original document and the final electronic record.

Examination forms and questionnaires will be reviewed and completed before data entry at the Zhongshan Ophthalmic Center. Range of data, frequency distribution and the consistency among related measurements are checked using data cleaning programs. The right eye of each student will be included in data analysis. If data for the right eye are not available, the left eye will be included in analysis.

## STATISTICAL ANALYSIS PLAN

Statistical analysis will be performed using Stata 13.1 software (StataCorp, College Station TX, USA). The distribution of baseline characteristics will be reported by use of mean (standard deviation) or median (interquartile range) for continuous variables, and frequency (percentage) for categorical variables. Baseline comparisons between study groups will be performed by using two-sample t test for continuous variables, and Chi-squared test for categorical variables. Mann-Whitney test will be used when dependent variables are not normally distributed. The differences between study groups in the change of SE and AL will be calculated using the logistic regression analysis. Cumulative incidence of myopia and their associated risk factors will be estimated using logistic regression. A two-sided P < 0.05 will considered statistically significant. Multiple imputation will be usd to handle missing data.

## STUDY MONITORING

At the start of the project, the Case Report Form, work flow, quality control procedures, electronic questionnaire, data entry methods, and follow-up management will be detailed in the training courses.

## PATIENT AND PUBLIC INVOLVEMENT

Participants are not invited to comment on the study design and aims or to interpret the findings, contribute to the writing or editing of this document.

## ETHICS AND DISSEMINATION

This project has obtained the ethics approval from the ethics committee of the Zhongshan Ophthalmic Center (number: 2019KYPJ171). The content of scientific research complies with the principles of the Declaration of Helsinki and its regulations applicable to research and human participants. The study investigators should protect participants’ privacy and are responsible for the confidentiality of all private information. The principal investigator is responsible for informing the ethics committee of any amendments to the protocol. The baseline and annual results will be published in peer-reviewed journals.

Clinical records and datasets will be kept at the Zhongshan Ophthalmic Center. Any public report of the results will not disclose personal information. Data will be de-identified before being passed to the study statisticians. Data will be kept in strict confidence and will only be assessed by the study investigators and authorized personnel. All data documents will be password protected, stored on a secure server, kept for at least 10 years and destroyed if no longer retention is required. If the project is modified, it will be exposed on Clinicaltrials.gov (Identifier: NCT04219228).

## DISCUSSION

It is estimated that the financial burden of myopia-related visual impairment reached $244 billion in 2015 globally.^18^ Since myopia generally develops during the school years, and tends to stabilize in adulthood, interventions to control the development of myopia need to be implemented during the school years.^1^ It is therefore imperative to explore the risk factors and predict the development of myopia among school-age children. In this current study, we will examine the association between environmental factors and myopia in urban and rural settings using subjective and objective data collection methods. Also, we will use neural learning algorithm to integrate ocular biological characteristics and environmental variables to establish an artificial intelligence model. In this way, we can effectively predict the occurrence and progression of myopia, and lay the foundation for individualized myopia prevention and control in the future.

## Data Availability

This article is a clinical study protocol and does not apply to this statement

## ACKNOWLEDGMENTS

We are grateful to Zhaoqing Education Bureau for providing eye examination database of students and all involved staff for their cooperation.

## DATA STATEMENT

Study datasets are currently unavailable.

## AUTHOR CONTRIBUTIONS

IM, YL, and Yingfeng Z conceived and designed the study. XC, GY, Yuxin Z, Yangfa Z, and Yingfeng Z wrote the draft. Yangfa Z, IM, and Yingfeng Z revised draft. LJ will lead the statistical analysis. Yingfeng Z and Yuxin Z will oversee data acquisition and implementation on site. All authors reviewed and approved the final manuscript.

## PATIENT AND PUBLIC INVOLVEMENT

Patients and/or the public were not involved in the design, or conduct, or reporting, or dissemination plans of this research.

